# Efficacy and safety of treating chronic nonspecific low back pain with radial extracorporeal shock wave therapy (rESWT), rESWT combined with celecoxib and eperisone (C+E) or C+E alone: a prospective, randomized trial

**DOI:** 10.1101/2020.11.28.20240119

**Authors:** Xuejiao Guo, Lin Li, Zhe Yan, Yunze Li, Zhiyou Peng, Yixin Yang, Yanfeng Zhang, Christoph Schmitz, Zhiying Feng

## Abstract

**Background:** To investigate whether respectively radial extracoporeal shock wave therapy (rESWT) or a combination of rESWT, Celecoxib and Eperisone (rESWT+C+E) are superior in reducing pain in subjects with chronic nonspecific low back pain (cnsLBP) compared to C+E alone (a standard treatment of this condition in China).

**Methods:** 140 subjects with cnsLBP were randomly allocated to rESWT (n=47), rESWT+C+E (n=45) or C+E alone (n=48) for four weeks. Outcome was evaluated using the Pain Self-Efficacy Questionnaire (PSEQ), Numerical Rating Scale (NRS), Oswestry Low Back Pain Disability Questionnaire and Patient Health Questionnaire 9, collected at one week (W1), W2, W3, W4 and W12 after beginning of the therapy.

**Results:** All scores showed a statistically significant improvement over time. The PSEQ and NRS scores showed a significant Time × Treatment effect. Subjects treated with rESWT had significantly lower mean NRS values than subjects treated with rESWT+C+E at W1 and W3, as well as than subjects treated with C+E alone at W3 and W4. No severe adverse events were observed.

**Conclusion:** rESWT may be superior to respectively rESWT+C+E or C+E alone in reducing pain in subjects with cnsLBP. Level of Evidence: Level I, prospective, randomized, active-controlled trial.

## 1. Introduction

Nonspecific low back pain (nsLBP) is defined as low back pain not attributable to a recognizable, known specific pathology [1, 2]. Nonspecific low back pain is common and affects people of all ages [2-4]. It is among the leading complaints bringing patients to physicians’ offices [2, 4]. The reported point prevalence of nsLBP is as high as 33 percent [2, 5], and its one-year prevalence is as high as 73 percent [2, 6]. Furthermore, its lifetime prevalence exceeds 70% in most industrialized countries [1], with an annual incidence of 15% to 20% in the USA [4]. In physically active adults not seeking medical attention, the annual incidence of clinically significant nsLBP (pain level, 4 or more on a 10-point scale) with functional impairment is approximately 10 to 15 percent [3]. In China, nsLBP has become one of the leading causes of disability-adjusted life-years in 2010, next to cardiovascular diseases, cancer and depression [7]. The prevalence of nsLBP was reported as 41% in Chinese adolescents, as well as a close relationship between nsLBP and self-reported academic pressure [8].

An alarming increase in the prevalence of chronic nsLBP (cnsLBP) has been observed in industrialized countries over the last years (note that different definitions of chronicity of nsLBP were proposed in the literature, mainly characterized by the duration of symptoms [9], e.g., more than seven weeks [10], more than three months [11-13] – which is used in the present study - or at least half the days in a 12-month period in a single or in multiple episodes [14]). For instance, in North Carolina (USA) the prevalence of chronic, impairing nsLBP increased from 3.9 percent in 1992 to 10.2 percent in 2006 [15]. Increases were seen in both men and women, and across all ages and racial and ethnic groups. In China, the years lived with disability showed a 44% increase from 1990 to 2010 [7].

Management of nsLBP is challenged by the problems that most back pain has no recognizable cause (>85%), an underlying systemic disease is rare, and most episodes of back pain are unpreventable [2-4]. Fortunately, acute nsLBP (lasting three to six weeks) usually resolves in several weeks, although recurrences are common and low-grade symptoms are often present years after an initial episode. Risk factors for the development of disabling chronic or persistent nsLBP include preexisting psychological distress, disputed compensation issues, other types of chronic pain and job dissatisfaction [3, 16, 17].

Recently the American College of Physicians issued a novel guideline for treating nonradicular low back pain [18]. According to this guideline acute and subacute nsLBP should be treated with superficial heat, massage, acupuncture and spinal manipulation, or with nonsteroidal anti-inflammatory drugs (NSAIDs) and skeletal muscle relaxants (SMRs), respectively. Chronic nsLBP should be treated with exercise, multidisciplinary rehabilitation, acupuncture, stress reduction, tai chi, yoga, motor control exercise, progressive relaxation, electromyography biofeedback, low-level laser therapy, operant therapy, cognitive behavioral therapy and spinal manipulation. In case of inadequate response to nonpharmacologic therapy, treatment with NSAIDs (first-line therapy) or tramadol or duloxetine (second-line therapy) should be considered [18].

In a recent consensus statement the Chinese Association for the Study of Pain has considered NSAIDs (including the selective Cox-2 inhibitor celecoxib), SMRs (including eperisone) and physiotherapy and rehabilitation (including exercise therapy, physiotherapy and psychotherapy) as first-line treatments of cnsLBP [19].

Unfortunately, few if any treatments have been proven effective for nsLBP in meta-analyses, including limited bed rest [20], physical activity and exercise [21], back schools [22], traction [23], massage [24], chiropractic [25, 26], radiofrequency denervation [27], paracetamol [28] and opioids [29]. Furthermore, earlier studies demonstrated that SMRs are effective in the management of nsLBP, but adverse effects require that these SMRs be used with caution [30]. Accordingly, the aforementioned guideline by the American College of Physicians [18] is only based on low- and moderate-quality evidence (c.f. also [31]).

During the last years a number of studies indicated that extracorporeal shock wave therapy (ESWT) may serve as an alternative in treatment of cnsLBP [32-42] (an overview on these studies is provided in Appendix A). However, in none of these studies ESWT was compared with NSAIDs and SMRs, let alone the investigation of potential advantages of the combination of ESWT with NSAIDs and SMRs over respectively ESWT or NSAIDs and SMRs alone. In consequence, these studies are of limited help to further corroborate or challenge the aforementioned guideline of the American College of Physicians [18] and the consensus statement by the Chinese Association for the Study of Pain [19].

Considering (i) the high incidence of cnsLBP, (ii) the limited evidence on which the aforementioned guideline of the American College of Physicians [18] and the consensus statement by the Chinese Association for the Study of Pain [19] are based and (iii) the limited relevance of all studies on treatment of cnsLBP with ESWT that were published so far [32-42], it was the aim of the present study to test the following hypotheses in a prospective, randomized, active-controlled trial: (i) treatment of cnsLBP with radial ESWT (rESWT) is safe; and (ii) both rESWT and the combination of rESWT, celecoxib and eperisone (rESWT+C+E) are superior to celecoxib and eperisone (C+E) alone in treatment of cnsLBP.

### 2. Experimental Section

This prospective, randomized, active-controlled trial was performed at the Department of Pain Medicine of the First Affiliated Hospital, Zhejiang University School of Medicine, Hangzhou, China (hereafter: “our department”). A total of n=152 subjects suffering from cnsLBP were assessed for eligibility to be enrolled in this study between October 2017 and March 2019. All subjects were from the city of Hangzhou and from other cities in Zhejiang province (China). Diagnosis was based on the subject’s history, physical examination at our department and X-rays of the lumbar spine. Subjects were considered for participation in the present study according to the inclusion and exclusion criteria summarized in Table 1.

**Table 1.** Inclusion and exclusion criteria of subjects with chronic nonspecific low back pain enrolled in the present study.

|  |
| --- |
| **Inclusion criteria** |
| Adults (both male and female) with nonspecific low back pain for more than three months |
| Age range: between 18 and 80 years |
| Willingness of the subject to participate in the study, and written informed consent signed and personally dated by the subject |
| Chronic nonspecific low back pain clinically diagnosed as repeated lumbar sourness and swelling pain or a chronic progressive process, accompanied by (i) X-ray examination to exclude lumbar vertebrate fractures, spondylolysis, spondylolisthesis and severe osteoporosis, and/or (ii) MRI with normal signal or low nucleus pulposus signal |
| No contraindications of rESWT or treatment with celecoxib and eperisone |
| **Exclusion criteria** |
| Children and teenagers below the age of 18 |
| Elderly aged >80 years old |
| No willingness of the subject to participate in the study, and/or written informed consent not signed and not personally dated by the subject |
| Previous spinal fracture or spinal surgery |
| Protrusion of a lumbar intervertebral disk, ankylosing spondylitis, scoliosis, lumbar spondylolisthesis and lumbar spondylolysis |
| Systemic disorders and psychiatric disorders |
| Contraindications of rESWT (pregnancy, presence of blood-clotting disorders (including local thrombosis), presence of local tumors, local bacterial and/or viral infections (including lumbar vertebral tuberculosis), treatment with oral anticoagulants and/or local corticosteroid applications in the period of six weeks before the first rESWT session (if applicable) |
| Contraindications of treatment with celecoxib and eperisone (allergy to celecoxib, eperisone or sulfonamides, presence of gastrointestinal bleeding or bleeding history, renal dysfunction and/or severe heart failure, and lactating women). |
| Participation in any other clinical trial in the period of 12 weeks before potential inclusion in the present study. |

### 2.1. Ethics committee approval

The present study was approved by the Ethics Committee of the First Affiliated Hospital, Zhejiang University School of Medicine, Hangzhou, China (no. 2017-515, dated 27/07/2017) and was carried out in accordance with the World Medical Association Declaration of Helsinki. Subjects were allowed to withdraw the free and informed consent term to participate in the study at any time.

The study has been registered with ClinicalTrials.gov (Identifier NCT03337607).

### 2.2. Procedures

Before randomization, a thorough explanation of the various options, as well as the potential risks, benefits and outcomes associated with the various options, took place. A total of 12 subjects assessed for eligibility were excluded because they did not meet the inclusion criteria (one subject) or the subjects chose to withdraw or declined to sign the consent form (eleven subjects). After having obtained written informed consent from each of the remaining n=140 subjects, they were randomly assigned to receive respectively rESWT (n=47), rESWT+C+E (n=45) or C+E alone (n=48).

Randomization was performed by a person uninvolved in the study at our department using a computerized random number generator. The results of randomization were kept in sealed opaque envelopes, thus keeping allocation concealed to both subjects and therapists until treatment started. Characteristics of included subjects at baseline are summarized in Table 2; individual localization of cnsLBP is displayed in Figure 1.

**Table 2.** Characteristics of included subjects at baseline (intention-to-treat population). a, duration of symptoms before enrollment into the present study. b, 0, no education; 1, pupil; 2, middle school; 3, higher education. c, 1, not married; 2, married; 3, divorced; 4, widow. *, Sjogren’s syndrome. Abbreviations: SD, standard deviation; BMI, body mass index; PSEQ, Pain Self-Efficacy Questionnaire; NRS, numerical rating scale; OLDPDQ, Oswestry Low Back Pain Disability Questionnaire; PHQ-9, Patient Health Questionnaire 9.

| Variable \ Group | rESWT (n=47) | rESWT+C+E (n=45) | C+E (n=48) |
| --- | --- | --- | --- |
| Age, years, median; mean (SD; min; max) | 34; 34.9 (8.7; 21; 58) | 34; 36.5 (10.8; 21; 64) | 32; 36.0 (11.2; 21; 70) |
| Woman, n (%) | 22 (46.8) | 28 (62.2) | 25 (52.1) |
| Body weight, kg, median; mean (SD; min; max) | 60; 62.3 (11.2; 41; 82) | 58; 60.8 (11.3; 40; 82) | 67; 64.4 (10.2; 42; 80) |
| Body height, cm, median; mean (SD; min; max) | 167; 166 (7.3; 153; 185) | 161; 163 (7.6; 148; 180) | 163; 165 (8.9; 130; 183) |
| BMI, kg/m<sup>2</sup>, median; mean (SD; min; max) | 22.6; 22.3 (3.0; 16.7; 28.3) | 22.3; 22.7 (3.2; 17.0; 31.2) | 23.8; 23.7 (3.7; 17.0; 41.4) |
| Duration<sup>a</sup>, months, median; mean (SD; min; max) | 12; 33.5 (45.8; 3; 240) | 24; 29.6 (30.2; 3; 120) | 12; 29.9 (49.1; 3; 240) |
| Education status<sup>b</sup> | 0; 1; 7; 39 | 0; 8; 7; 30 | 1; 5; 15; 27 |
| Marriage status<sup>c</sup> | 7, 39, 1, 0 | 9, 36, 0, 0 | 7, 40, 1, 0 |
| Comorbidities, n (diagnosis) | 1 (diabetes) | 0 | 1* |
| PSEQ score, median; mean (SD; min; max) | 55; 51.3 (8.5; 26; 60) | 50; 46.8 (12.0; 8; 60) | 53.5; 48.8 (10.9; 17; 60) |
| NRS score, median; mean (SD; min; max) | 4; 4.2 (1.2; 1; 6) | 4; 4.4 (1.4; 1; 7) | 4; 4.2 (1.5; 2; 8) |
| OLDPDQ score, median; mean (SD; min; max) | 14.0; 16.8 (11.4; 2; 66) | 20.0; 21.6 (12.8; 4; 68) | 18.0; 18.9 (10.4; 0; 50) |
| PHQ-9 score, median, mean (SD; min; max) | 3; 4.6 (4.1; 0; 14) | 5; 6.2 (5.1; 0; 22) | 6; 6.9 (4.7; 0; 19) |

**Figure 1.**
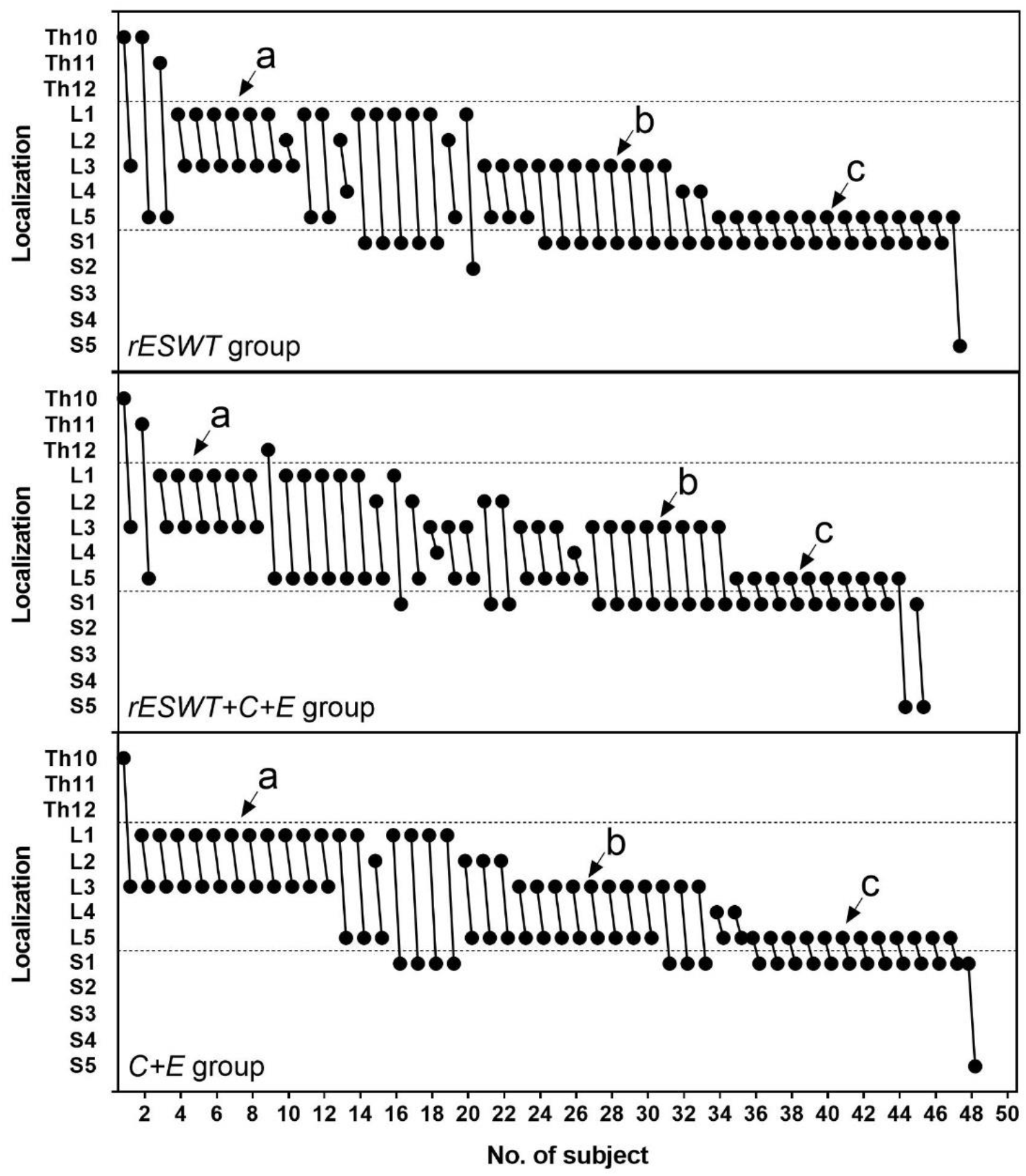
Individual localization of chronic nonspecific low back pain reported by the subjects enrolled in the present study. The arrows exemplarily indicate subjects who reported pain from lumbar segment L1 to L3 (a), from L3 to sacral segment S1 (b) or from L5 to S1 (c), respectively.

Subjects in the *rESWT* group received rESWT using the Swiss DolorClast device (Electro Medical Systems, Nyon, Switzerland) and the EvoBlue handpiece. Each subject received four rESWT sessions (one session per week). Each session consisted of a total of 4000 radial extracorporeal shock waves (rESWs) (1000 rESWs each applied to the left and the right paravertebrate muscles from the third lumbar segment (L3) to the first sacral segment (S1) using the 36-mm applicator, plus 1000 rESWs each applied to the left and the right sacroiliacal joint using the 15-mm convex applicator, in prone position of the subject). The air pressure of the rESWT device (and, thus, the energy flux density (EFD) of the applied rESWs) was gradually increased during the first 200 rESWs each until the maximum discomfort the subject could tolerate was reached, followed by 800 rESWs at this air pressure / EFD. The rESWs were applied at a frequency of 15 Hz. No anesthesia or analgesic drugs were applied during the rESWT sessions.

Subjects in the *rESWT+C+E* group received rESWT as the subjects in the *rESWT* group plus celecoxib (1 × 200 mg per day for moderate pain (Numerical Rating Scale (NRS) score 4-6) or 2 × 200 mg per day (NRS score 7-10)) and eperisone (3 × 50 mg per day) for four weeks.

Subjects in the *C+E* group received celecoxib and eperisone as the subjects in the *rESWT+C+E* group, but no rESWT.

In addition, all subjects were advised to perform simplified, safe core stability training and flexion / relaxation training at home, which was mainly based on the contraction of the lumbar muscles, under the guidance of a unified rehabilitation training video (two training sessions per week; each training session lasting for approximately 20 minutes; training for the entire duration of the study).

### 2.3. Outcome measures

The primary outcome measure was change in the Pain Self-Efficacy Questionnaire (PSEQ) score [43]. Specifically, subjects were asked to rate how confident they were at the time of examination despite the presence of their pain in performing the activities described in Table 3, listed by selecting a number on a 7 point scale where 0 equals “not at all confident” and 6 equals “completely confident”.

**Table 3.** Activities assessed by the Pain Self-Efficacy Questionnaire (PSEQ) [43].

| No. | Activity |
| --- | --- |
| 1 | I can enjoy things, despite the pain. |
| 2 | I can do most of the household chores (e.g. tidying-up, washing dishes, etc.), despite the pain. |
| 3 | I can socialize with my friends or family members as often as I used to do, despite the pain. |
| 4 | I can cope with my pain in most situations. |
| 5 | I can do some form of work, despite the pain (“work” includes housework, paid and unpaid work). |
| 6 | I can still do many of the things I enjoy doing, such as hobbies or leisure activity, despite the pain. |
| 7 | I can cope with my pain without additional medication (next to study treatment). |
| 8 | I can still accomplish most of my goals in life, despite the pain. |
| 9 | I can live a normal lifestyle, despite the pain. |
| 10 | I can gradually become more active, despite the pain. |

Scores on the PSEQ may range from 0 to 60, with higher scores indicating stronger selfefficacy beliefs. A point score change of 11 points for the PSEQ score corresponds to the minimal clinically important difference (MCID), which is described in the literature as the smallest difference that subjects and clinicians perceive to be worthwhile when treating chronic nonspecific low back pain [44]. According to the latter study the PSEQ score is responsive to clinically important change over time. The PSEQ score was collected at baseline (BL) and at all follow-up examinations, i.e., one week (W1), two weeks (W2), three weeks (W3), four weeks (W4) and twelve weeks (W12) after beginning of the treatments.

In the protocol of the present study treatment success was defined as individual improvement of the PSEQ score by more than 20 points at W12.

Secondary clinical outcome measures were changes in the Numerical Rating Scale (NRS) score (subjects were asked to rate their pain intensity on an 11-point scale where 0 indicated no pain at all and 10 indicated worst imaginable pain), the Oswestry Low Back Pain Disability Questionnaire (OLDPDQ) score [45, 46] and the Patient Health Questionnaire 9 (PHQ-9) score [47]. The NRS, OLDPDQ and PHQ-9 scores were collected at BL, W1, W2, W3, W4 and W12.

Complications, adverse events and complaints during treatment were documented.

### 2.4. Blinding

The study protocol did not allow to blind the subjects and therapists who performed the treatments. On the other hand, the study investigators and evaluators were blinded for the entire duration of the study. Specifically, the study investigators and evaluators did not have access to the subjects’ treatment records, including subject allocation or the allocation sequence, until all subjects had completed the last follow-up examination.

### 2.5. Drop-outs and loss to follow-up

The flow of subjects in the present study according to the Consolidated Standards of Reporting Trials (CONSORT) statement [48] is shown in Figure 2. All subjects received treatment as allocated. None of the 47 subjects in the *rESWT* group, four out of the 45 subjects in the *rESWT+C+E* group and four out of the 48 subjects in the *C+E* group were lost to final follow-up at W12, resulting in full analysis of respectively 47/47 (100%) of the subjects in the *rESWT* group, 41/45 (91.1%) of the subjects in the *rESWT+C+E* group and 44/48 (91.7%) of the subjects in the *C+E* group who were randomized.

**Figure 2.**
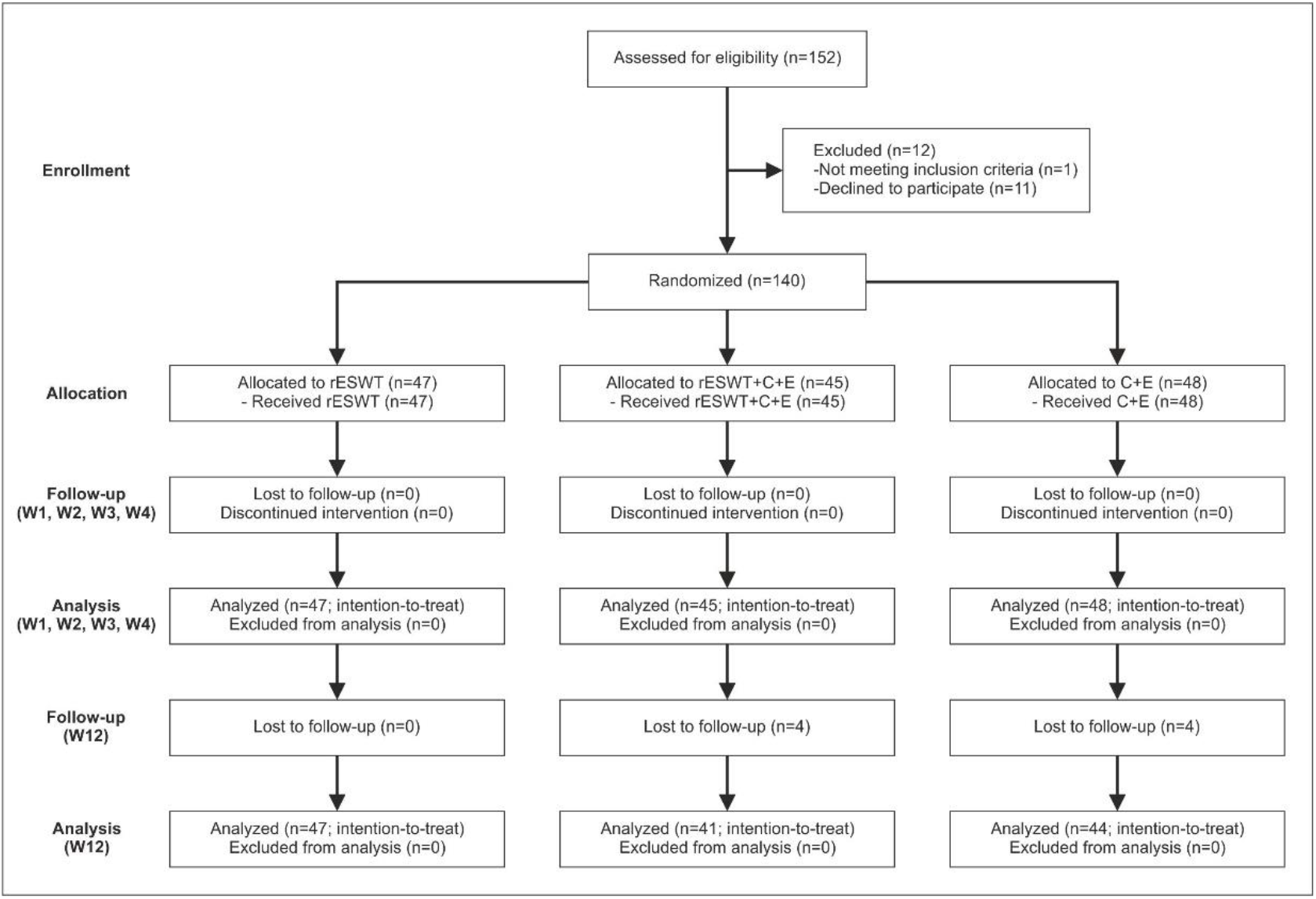
Flow of subjects in the present study according to the Consolidated Standards of Reporting Trials (CONSORT) statement [48].

### 2.6. Power analysis

In all studies on ESWT for nsLBP that were published at the time of registering the present study with ClinicalTrials.gov [32-35] no definition of treatment success was provided, no Power analysis was reported and the PSEQ score was not used to assess clinical outcome (c.f. Appendix 1). Therefore, these studies were only of very limited use for the purpose of performing a Power analysis of the present study.

Based on anecdotal evidence from several therapists in Europe and Latin America who had used rESWT for treating cnsLBP for more than a decade we hypothesized that treatment of cnsLBP with rESWT as described above will result in a success rate of approximately 70%, and the combination of rESWT+C+E in a success rate of approximately 85%. Furthermore, based on our own experience we hypothesized that treatment of cnsLBP with C+E as described above will result in a success rate of only approximately 40%.

Considering a two-sided significance level of 95%, power of 0.8 and equal samples, the power analysis retrieved a minimum number of respectively n=44 (according to [49]) or n=42 (according to [50]) subjects per group to be enrolled in the present study. The power analysis was performed with the online tool, Open Source Epidemiologic Statistics for Public Health [51].

### 2.7. Statistical analysis

Statistical analysis was performed on an intention-to-treat basis using the Last Observation Carried Forward Approach [52].

Mean and standard deviation were calculated for all investigated variables. The D’Agostino and Pearson omnibus normality test was used to determine whether the distribution of the investigated variables of the subjects in the different treatment groups were consistent with a Gaussian distribution. According to [53] no significance tests of baseline differences were performed.

Treatment-related differences in mean PSEQ scores, mean NRS scores, mean OLDPDQ scores and mean PHQ-9 scores between the groups were tested with two-way repeated measures ANOVA, with the different follow-up times (BL, W1, W2, W3, W4 and W12) as within-subject factor and the different groups as between-subject factor. Comparisons between groups were performed using Bonferroni’s multiple comparisons test.

In all analyses an effect was considered statistically significant if its associated P value was smaller than 0.05. Calculations were performed using GraphPad Prism (Version 9.0.0; GraphPad software, San Diego, CA, USA).

## 3. Results

No apparent differences between the groups were observed at baseline except for the mean PHQ-9 score (Table 2).

All investigated variables showed substantial interindividual variation and a statistically significant improvement over time (Figure 3 and Table 4).

**Table 4.** Results (p values) of the statistical analysis of the data shown in Figure 3 using two-way repeated measures ANOVA and Bonferroni’s multiple comparisons (m.c.) test. P values < 0.05 are given boldface.

| ANOVA: source of variation |  | PSEQ score | NRS score | OLBPDQ score | PHQ-9 score |
| --- | --- | --- | --- | --- | --- |
| Time |  | <0.001 | <0.001 | <0.001 | <0.001 |
| Treatment |  | 0.339 | 0.067 | 0.367 | **0.027** |
| Time × Treatment |  | **0.044** | **0.016** | 0.538 | 0.423 |
| Subject |  | <0.001 | <0.001 | <0.001 | <0.001 |
| Bonferroni's m.c. test |  | PSEQ score | NRS score | OLBPDQ score | PHQ-9 score |
| W0 | rESWT vs rESWT+C+E | 0.063 | >0.999 | 0.091 | 0.217 |
|  | rESWT vs C+E | 0.561 | >0.999 | >0.999 | **0.027** |
|  | rESWT+C+E vs C+E | 0.920 | >0.999 | 0.646 | >0.999 |
| W1 | rESWT vs rESWT+C+E | 0.199 | **0.027** | 0.370 | 0.850 |
|  | rESWT vs C+E | 0.921 | 0.540 | >0.999 | **0.011** |
|  | rESWT+C+E vs C+E | >0.999 | 0.579 | >0.999 | 0.219 |
| W2 | rESWT vs rESWT+C+E | >0.999 | 0.130 | 0.830 | >0.999 |
|  | rESWT vs C+E | 0.331 | 0.163 | >0.999 | **0.010** |
|  | rESWT+C+E vs C+E | 0.443 | >0.999 | 0.786 | 0.126 |
| W3 | rESWT vs rESWT+C+E | >0.999 | **0.029** | 0.669 | >0.999 |
|  | rESWT vs C+E | 0.232 | **0.010** | >0.999 | 0.082 |
|  | rESWT+C+E vs C+E | 0.921 | >0.999 | >0.999 | 0.370 |
| W4 | rESWT vs rESWT+C+E | >0.999 | 0.077 | 0.863 | 0.755 |
|  | rESWT vs C+E | >0.999 | **0.046** | >0.999 | **0.043** |
|  | rESWT+C+E vs C+E | >0.999 | >0.999 | >0.999 | 0.608 |
| W12 | rESWT vs rESWT+C+E | >0.999 | >0.999 | >0.999 | >0.999 |
|  | rESWT vs C+E | >0.999 | >0.999 | >0.999 | 0.324 |
|  | rESWT+C+E vs C+E | >0.999 | >0.999 | >0.999 | >0.999 |

**Figure 3.**
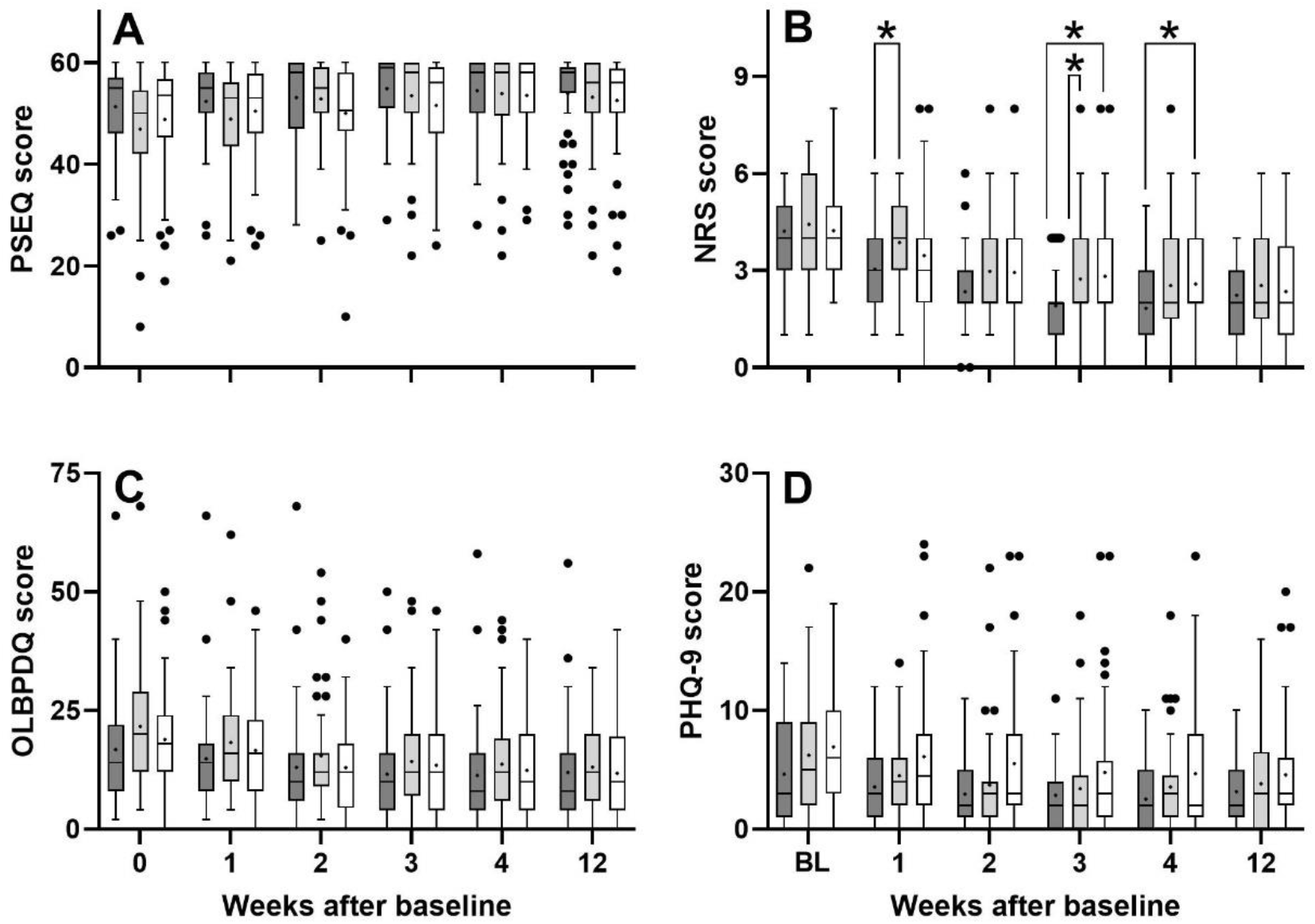
Tukey boxplots of (A) PSEQ score, (B) NRS score, (C) OLBPDQ score and (D) PHQ-9 score of subjects suffering from chronic nonspecific low back pain who were treated with respectively rESWT (dark gray bars), rESWT+C+E (light gray bars) or C+E alone (open bars) at baseline (X=0) and different follow-up times. Results of statistical analysis using Bonferroni’s multiple comparison test are indicated (*, p < 0.05).

The PSEQ score showed a statistically significant Time × Treatment effect, with no significant difference between groups at any investigated time (Table 4). The number of subjects with treatment success as defined in the protocol of the present study at W1 / W2 / W3 / W4 /W12 after baseline was 1 / 0 / 1 / 2 / 2 among the subjects in the *rESWT* group, 0 / 3 / 4 / 4 / 4 among the subjects in the *rESWT+C+E* group and 2 / 3 / 2 / 4 / 4 among the subjects in the *C+E* group. On the other hand, the number of subjects who could not reach treatment success as defined in the protocol of the present study because they had a PSEQ score > 40 at baseline was 41 (87.2%) among the subjects in the *rESWT* group, 35 (77.8%) among the subjects in the *rESWT+C+E* group and 39 (81.3%) among the subjects in the *C+E* group. Furthermore, the number of subjects who could not reach the MCID of 11 points because they had a PSEQ score > 49 at baseline was 31 (66.0%) among the subjects in the *rESWT* group, 24 (53.3%) among the subjects in the *rESWT+C+E* group and 29 (60.4%) among the subjects in the *C+E* group.

The NRS score also showed a statistically significant Time × Treatment effect (Figure 3 and Tables 4 and 5). Bonferroni’s multiple comparisons test showed no statistically significant difference between the groups at baseline (rESWT: 4.2 ± 1.2; rESWT+C+E: 4.4 ± 1.4; C+E: 4.2 ± 1.5) but between the subjects in the *rESWT* group and those in the *rESWT+C+E* group at W1 after baseline (3.0 ± 1.2 vs. 3.9 ± 1.4) and W3 after baseline (1.9 ± 1.3 vs. 2.7 ± 1.4), as well as between the subjects in the *rESWT* group and those in the *C+E* group at W3 after baseline (1.9 ± 1.3 vs. 2.8 ± 1.9) and W4 after baseline (1.8 ± 1.4 vs. 2.6 ± 1.7).

**Table 5.** Mean ± standard deviation of the NRS score before and at different times after baseline in the present study. Abbreviations: W1 / W2 / W3 / W4 / W12, one / two / three / four / twelve weeks after baseline; C, change compared to baseline.

| Treatment | rESWT |  | rESWT+C+E |  | C+E |  |
| --- | --- | --- | --- | --- | --- | --- |
| Baseline | 4.2 ± 1.2 | | 4.4 ± 1.4 | | 4.2 ± 1.5 | |
| W1 / C | 3.0 ± 1.2 | -27.8% | 3.9 ± 1.4 | -12.6% | 3.5 ± 1.9 | -18.2% |
| W2 / C | 2.3 ± 1.2 | -44.5% | 3.0 ± 1.4 | -32.7% | 2.9 ± 1.8 | -30.5% |
| W3 / C | 1.9 ± 1.3 | -54.5% | 2.7 ± 1.4 | -38.2% | 2.8 ± 1.9 | -33.2% |
| W4 / C | 1.8 ± 1.4 | -56.6% | 2.5 ± 1.6 | -42.7% | 2.6 ± 1.7 | -38.9% |
| W12 / C | 2.2 ± 1.3 | -47.7% | 2.5 ± 1.6 | -42.7% | 2.4 ± 1.6 | -44.3% |

The subjects in the *rESWT* group had statistically significantly lower mean PHQ-9 scores than the subjects in the *C+E* group at baseline (4.6 ± 4.1 vs. 6.9 ± 4.7), W1 after baseline (4.6 ± 4.4 vs. 6.9 ± 4.7), W2 after baseline (3.6 ± 3.0 vs. 6.1 ± 5.5) and W4 after baseline (2.9 ± 2.9 vs. 4.8 ± 5.4).

No severe adverse events were observed during the present study.

## 4. Discussion

This is the first study that compared the efficacy and safety of rESWT, rESWT+C+E and C+E alone in treatment of cnsLBP. Several conclusions can be drawn from the results of the present study, some of which appear of relevance mostly to China, whereas others are of relevance to therapists worldwide.

A key finding of the present study was that the PSEQ score [43] turned out to be unsuitable for evaluating treatment success in the present study. This was due to the fact that 115 out of the 140 subjects (82.1%) investigated in the present study had a PSEQ score of 41 or higher at baseline and, thus, could not reach treatment success as defined in the protocol of the present study. Furthermore, 83 out of the 140 subjects (59.3%) had a PSEQ score of 50 or higher at baseline and, thus, could not reach a point score change of 11 points which has been defined in the literature as MCID of the PSEQ score when treating cnsLBP [44]. In this regard it is crucial to bear in mind that the subjects investigated in the present study were not specifically selected. Rather, they are representative for all subjects who seek medical treatment of cnsLBP at our department. It is quite possible that in other countries, a substantial amount of these subjects would have consulted their family doctor before visiting a department of pain medicine at a hospital affiliated with a university school of medicine. However, a family doctor system does not exist in China.

Until now the PSEQ score has not been used in any study listed in PubMed on treatment of cnsLBP performed at a department of pain medicine in China. One study on this topic authored by Chinese authors has been listed in PubMed so far [54]. The subjects investigated in this study were recruited from the Rehabilitation Clinic of The Hong Kong Polytechnic University and were treated with respectively physiotherapy (Group 1) or physiotherapy + smartphone-based remote self-management (Group 2). The mean PSEQ scores at baseline in [54] (Group 1: 34.3 ± 8.0; Group 2: 38.6 ± 8.5; mean ± SD) were lower than the mean PSEQ score of the subjects investigated in the present study (49.0 ± 10.6). However, there are a number of substantial shortcomings in [54] which renders this study insufficient for contributing to evaluating the mean PSEQ score of subjects with cnsLBP who seek treatment at a department of pain medicine in China: (i) the number of subjects investigated in [54] was very small (Group 1: n=3; Group 2: n=5) and, thus, the total number of subjects investigated in [54] (n=8) was less than 20% of the number of subjects in each group investigated in the present study; and (ii) the only inclusion criteria reported in [54] were nonspecific low back pain due to musculoskeletal origins, access to a mobile phone and the ability to perform a brief exercise during regular working hours; and the only exclusion criterion reported in [54] was history of receiving major surgery. This is quite different to the inclusion and exclusion criteria used in the present study (Table 1).

Our finding of efficacy of rESWT for cnsLBP assessed using the NRS score (Figure 3B) is in line with several reports in the literature on both rESWT [32, 33, 38, 39] and focused ESWT (fESWT) [34, 41, 42] (c.f. Appendix A). On the other hand, there was no advantage in combining rESWT with C+E over respectively rESWT or C+E alone in treatment of cnsLBP in the present study (to our knowledge studies on C+E alone in treatment of cnsLBP have not been published so far). Rather, rESWT alone was superior to rESWT+C+E in pain relief (assessed using the NRS score) at W1 and W3 after baseline, and superior to C+E alone at W3 and W4 after baseline (Figure 3B). No other study on rESWT or fESWT for cnsLBP published so far (c.f. Appendix A) has addressed this important question about potential synergistic effects of combining rESWT or fESWT with pharmacological treatments in the management of cnsLBP.

With over 48.000 randomized controlled trials (RCTs), systematic reviews and clinical practice guidelines the PEDro database [55] is currently the largest independent database in the field of physical and rehabilitation medicine. All RCTs listed in the PEDro database are independently assessed for quality; all but two of the PEDro scale items are based on the Delphi list [56]. Currently, there are 1877 studies on low back pain listed in the PEDro database. Of note, only three out of these 1877 studies (0.16%) compared the efficacy of non-pharmacological, non-surgical treatments with pharmacological treatments and the combination thereof in the management of cnsLBP [57-59] (summarized in Appendix B). One study on acute low back pain [57] compared spinal manipulation (two to three treatment sessions per week for up to four weeks) with paracetamol (4 × 1g/day for up to four weeks) and the combination thereof and found no statistically significant reduction in the number of days to recovery of any of these treatments compared with placebo spinal manipulation and placebo drug treatment. Another study that comprised subjects suffering from respectively acute low back pain, migraine or ankle sprain [58] compared a single session of acupuncture with pharmacotherapy (different protocols; summarized in Appendix B) and the combination thereof and found that all of these strategies reduced mean pain by approximately 24% one hour after baseline (individual outcome for the different indications and longer follow-up times were not reported in [58]).

The third of these studies listed in the PEDro database [59] compared acupuncture (two treatment sessions per week for five weeks) with baclofen (a centrally acting skeletal muscle relaxant; [60]) (2 × 15 mg/day for five weeks) and the combination thereof in treatment of cnsLBP. The authors of this study found that both acupuncture and the combination of acupuncture and baclofen statistically significantly reduced the mean NRS score at both W5 and W10 after baseline (acupuncture: reduction by 30.2% at W5 after baseline and 21.9% at W10 after baseline; acupuncture + baclofen: reduction by 38.5% at W5 after baseline and 27.7% at W10 after baseline) but not baclofen alone. Furthermore, the combination of acupuncture and baclofen was statistically significantly more effective than acupuncture alone at W10 after baseline [59]. The percentage improvement in mean NRS score after treatment with acupuncture and baclofen at W5 / W10 after baseline in [59] was 32% / 42% lower than after treatment with rESWT at W4 / W12 after baseline in the present study. This may indicate that rESWT is superior to acupuncture in treating cnsLBP, which should be addressed in future studies (acupuncture was not used as control treatment in any of the studies on rESWT and fESWT in treatment of cnsLBP listed in Appendix A).

The question why in the present study treatment of cnsLBP with rESWT was superior to treatment with rESWT+C+E or C+E alone, whereas in [59] treatment with acupuncture + baclofen was superior to treatment with either acupuncture or balcofen alone, cannot be fully answered at this time. Although no study has systematically addressed similarities and differences in cellular and molecular mechanisms of action between extracorporeal shock waves (ESWs) and acupuncture, it is reasonable to hypothesize that these mechanisms are not the same. To make matters worse, the mode of action of eperisone (used in the present study) is different from the mode of action of baclofen (used in [59]). In cnsLBP ESWT most probably acts mainly via reduction of substance P and calcitonin gene-related peptide (CGRP) in C nerve fibers [61, 62]. Substance P nerve fibers were demonstrated within subchondral bone of degenerative lumbar facet joints in humans and hypothesized being involved in the etiology of low back pain [63]. Both substance P and CGRP were found in human cervical facet joint capsules [64]; substance P was also found in the lumbar facet joint capsule of rabbits [65] (c.f. also [66]). The high amount of substance P in active trigger points [67] as well as the ability of ESWs to impair acetylcholine receptors at the neuromuscular junction [68] and to increase the expression of lubricin in tendons and septa [69] (which improves the gliding ability of tendons and septa [70]) are additional factors that may contribute to the beneficial effects of ESWT in the management of cnsLBP.

It is well known that effects of ESWs on nociceptors can be blocked by local anesthesia [71] and, thus, treatment of tendinopathies with ESWT without local anesthesia is more effective than with local anesthesia [72, 73]. Eperisone is a centrally acting skeletal muscle relaxant which directly acts on motor nerves to hyperpolarize the action potential, resulting in reduction of nerve sensitivity and conduction of the muscle spindle contraction [74]. Besides this, eperisone has an analgesic effect, which apparently is mediated by inhibition of the release of substance P [75]. Accordingly, one may hypothesize that eperisone at least partially blocked the action of ESWs on substance P nerve fibers in the present study, which may explain why rESWT was superior to rESWT+C+E in treatment of cnsLBP in the present study. This phenomenon should be addressed in future studies.

Finally it should be mentioned that it remains unknown why despite randomization the subjects in the *C+E* group had a statistically significantly higher mean PHQ-9 score at baseline than the subjects in the *rESWT* group. However, according to [53] no adjustment of this subjects imbalance was performed (by, e.g., some form of multiple regression analysis). In any case, the key findings of the present study (rESWT is effective and safe in treatment of cnsLBP, and may be superior to rESWT+C+E) are independent of this subjects imbalance, because the mean PHQ-9 scores at baseline of the subjects in the *rESWT* group and those in the *rESWT+C+E* group were not statistically significantly different.

The present study has a number of limitations. One limitation is the relatively high mean PSEQ score of the subjects at baseline, which caused that the majority of subjects could neither reach a point score change of 20 points which was defined as treatment success in the protocol of the present study, nor a point score change of 11 points which has been defined in the literature as MCID of the PSEQ score when treating cnsLBP [44]. As outlined in detail above, this was due to the fact that the subjects investigated in the present study were not specifically selected but were representative for all subjects who seek medical treatment of cnsLBP at our department (the latter is representative for departments of pain medicine at hospitals affiliated with university schools of medicine in China). On the other hand, a post-hoc subgroup analysis of those subjects in the present study with a PSEQ score < 50 (i.e., those subjects who could have reached the MCID of the PSEQ score when treating cnsLBP) indicated that the present study would have come to similar conclusions if subjects would have been preselected (Appendix C). Another limitation is that only a single pharmacological treatment was tested in the present study. Accordingly, the question could not be answered whether any combination of rESWT with a pharmacological treatment (NSAIDs and SMRs) could be superior to rESWT alone in treatment of cnsLBP. This should be addressed in future studies with a different skeletal muscle relaxant than eperisone. Finally, the protocol of the present study did not allow to finally answer the question whether rESWT is non-inferior or even superior to rESWT+C+E or C+E alone. However, considering the potential negative interaction between ESWs and eperisome in treatment of cnsLBP this limitation appears irrelevant. Future studies on this topic will most probably not consider eperisome anymore in related combination therapies.

## 5. Conclusions

The present study suggests that the use of rESWT in subjects with cnsLBP who seek medical treatment at a department of pain medicine in China is safe and effective, leading to a significant reduction in pain, without adverse events. In this regard the present study corroborates similar findings from other parts of the world. Particularly in China clinicians should consider rESWT as a non-pharmacological alternative to celecoxib and eperisone in the management of cnsLBP. Based on apparently opposite effects on substance P nerve fibers the combination of rESWT and eperisone should not be considered anymore. Future studies should address whether the combination of rESWT with celexocib and/or other SMRs (including baclofen) is superior to rESWT or pharmacological treatment alone.

## Data Availability

All data are available upon request.

## Author Contributions

Conceptualization, X.J., C.S. and Z.F.; methodology, X.J, L.L, C.S. and Z.F.; software,Y.Z., Y.L and C.S.; validation, Y.Z., Y.L and Z.F.; formal analysis, C.S.; investigation,Y.Z.; resources, X.J, L.L.; data curation, Z.Y and Z.P.; writing—original draft preparation, C.S.; writing—review and editing, Y.Y and Z.F.; visualization, C.S.; supervision, Z.F.; project administration, C.S. and Z.F.; funding acquisition, not applicable. All authors have read and agreed to the published version of the manuscript.

## Funding

This research received no external funding.

### Acknowledgments

We express our gratitude to all the subjects with chronic nonspecific low back pain who were treated at the Department of Pain Medicine, The First Affiliated Hospital, Zhejiang University School of Medicine, Hangzhou, China between between October 2017 and March 2019, and whose data were considered in the present study. We also thank everyone at our department for their contribution to the study operations.

### Appendix A

Studies on radial extracorporeal shock wave therapy (rESWT) and focused extracorporeal shock wave therapy (fESWT) for nonspecific low back pain (LBP) that were published so far.

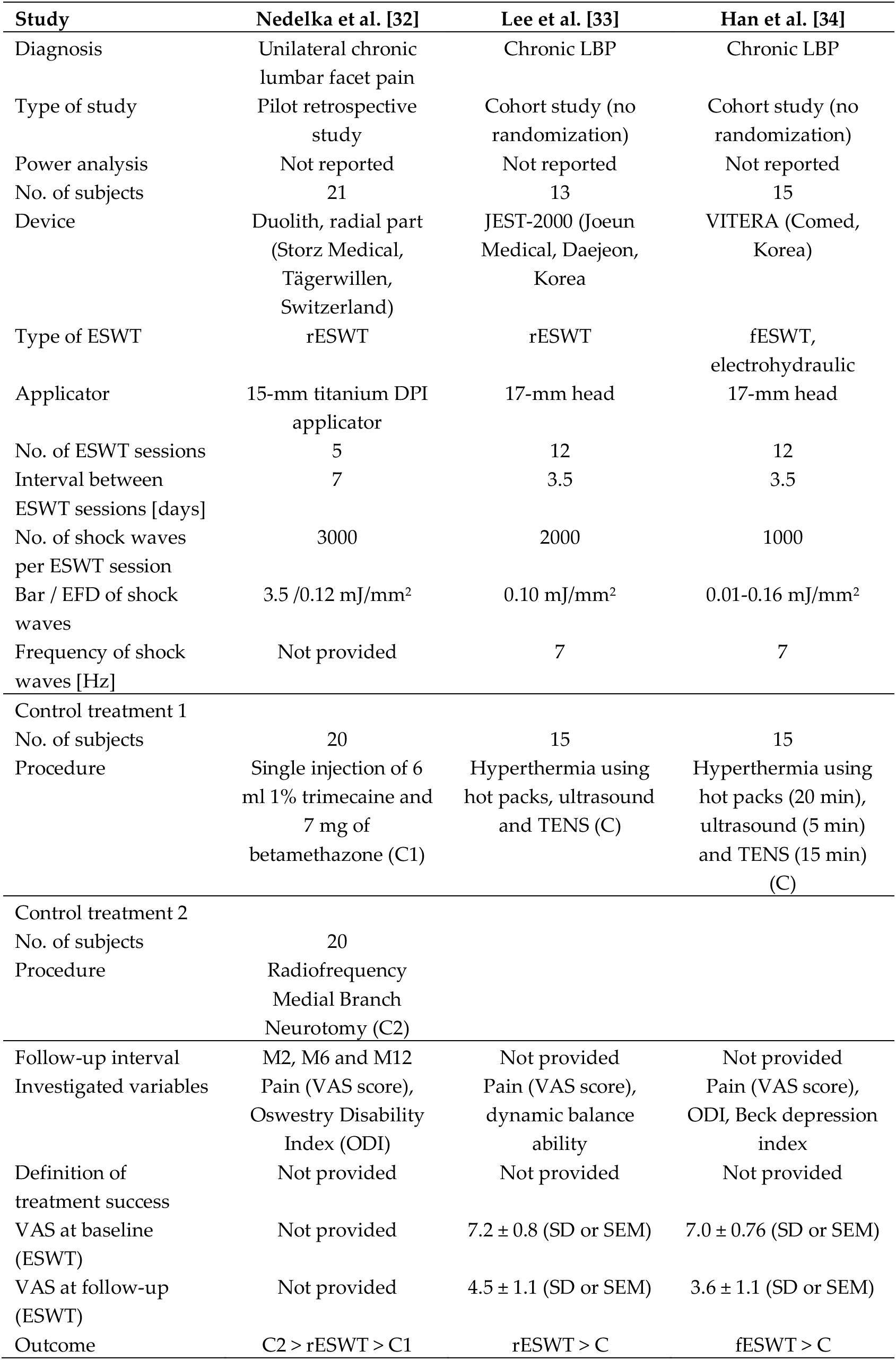
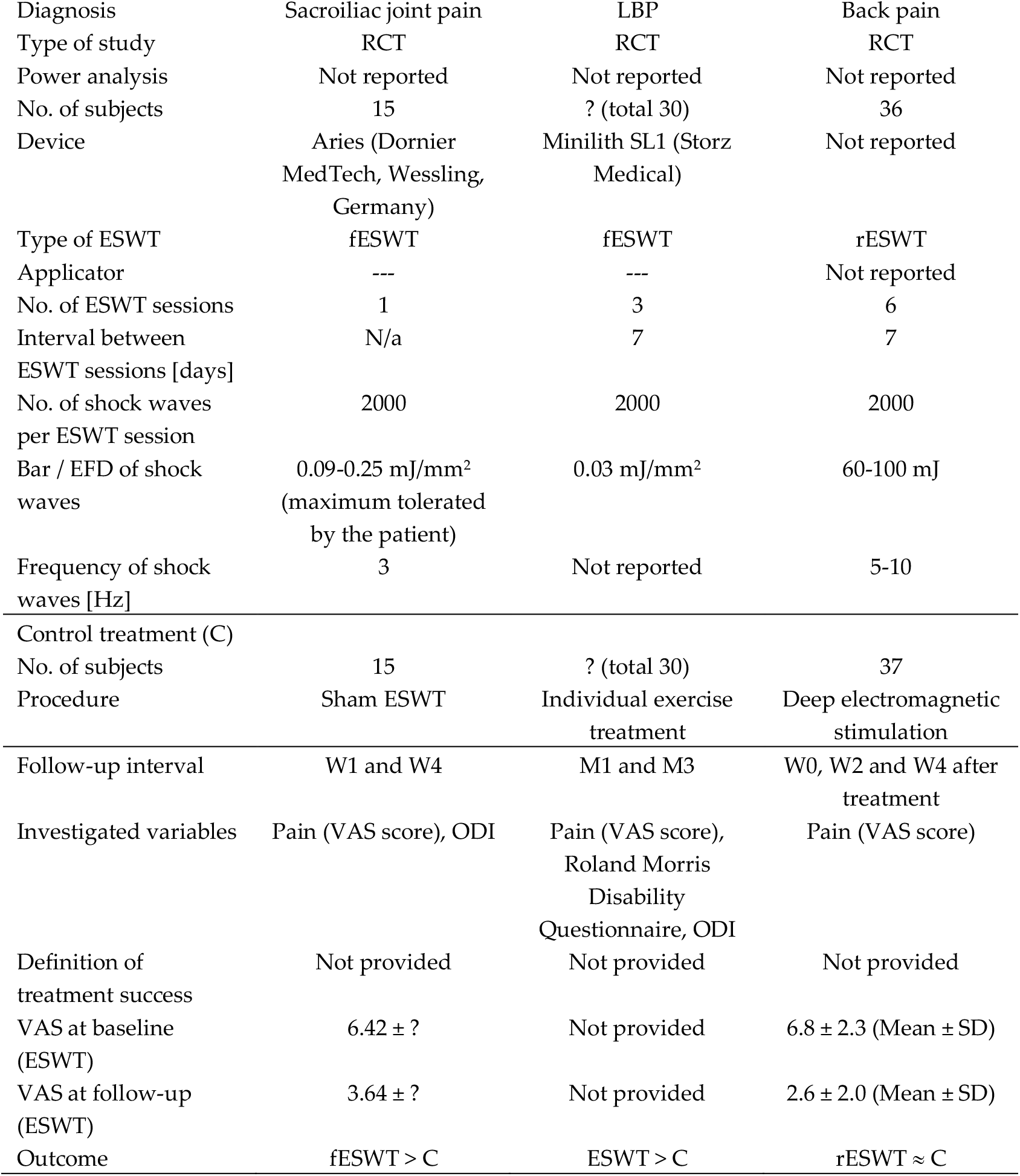
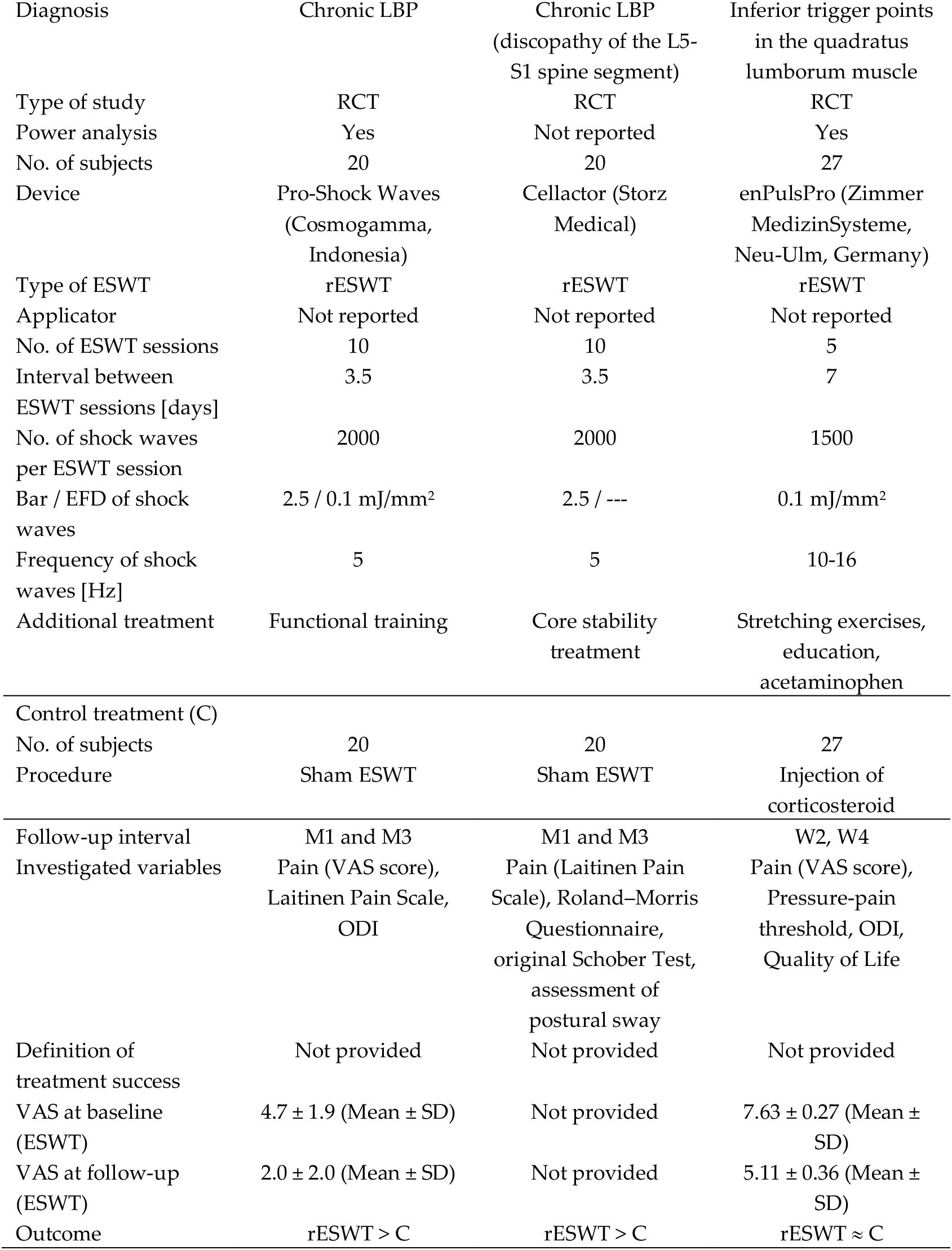
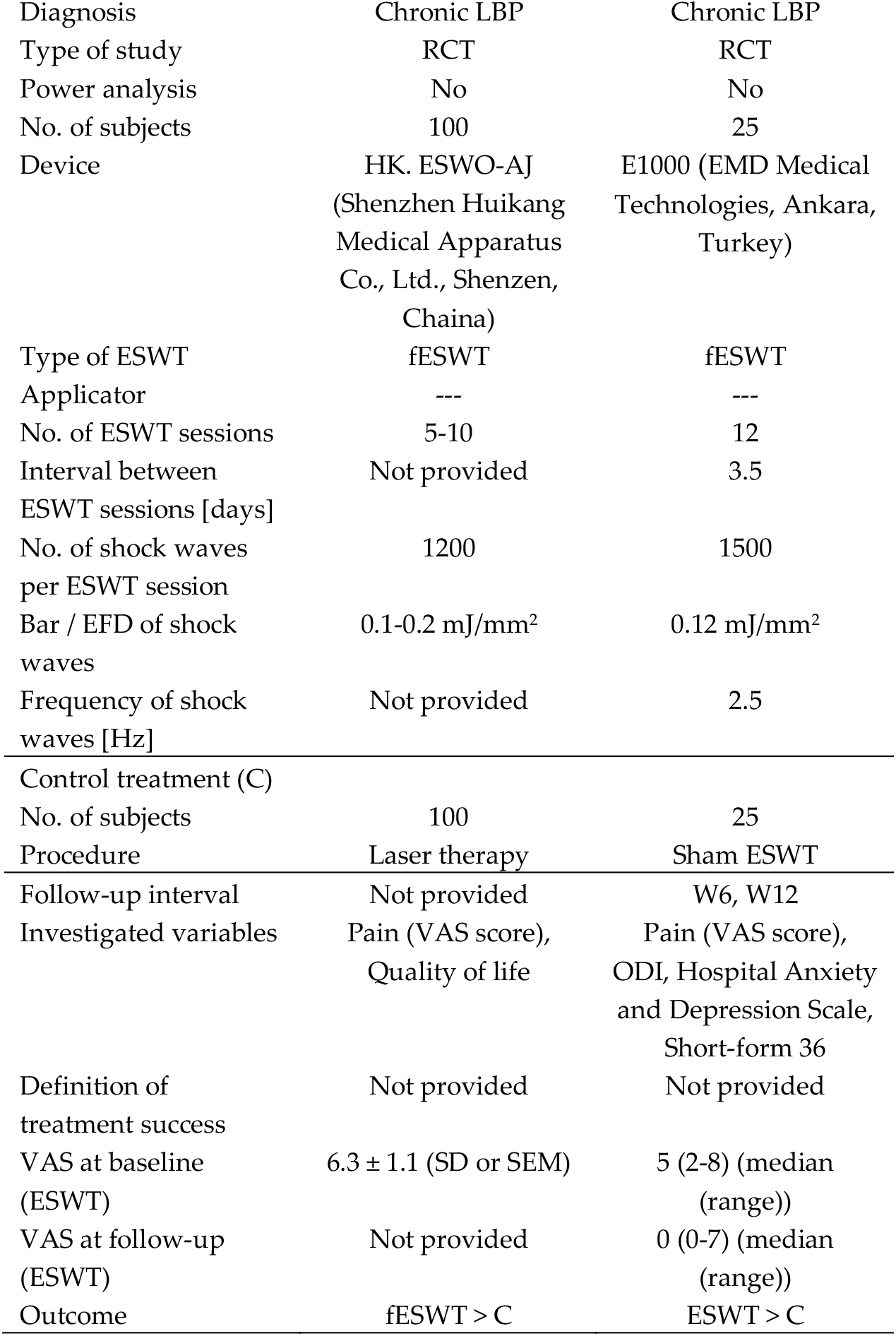

| Study | Nedelka et al. [32] | Lee et al. [33] | Han et al. [34] |
| --- | --- | --- | --- |
| Diagnosis | Unilateral chronic lumbar facet pain | Chronic LBP | Chronic LBP |
| Type of study | Pilot retrospective study | Cohort study (no randomization) | Cohort study (no randomization) |
| Power analysis | Not reported | Not reported | Not reported |
| No. of subjects | 21 | 13 | 15 |
| Device | Duolith, radial part (Storz Medical, Tägerwillen, Switzerland) | JEST-2000 (Joeun Medical, Daejeon, Korea) | VITERA (Comed, Korea) |
| Type of ESWT | rESWT | rESWT | fESWT, electrohydraulic |
| Applicator | 15-mm titanium DPI applicator | 17-mm head | 17-mm head |
| No. of ESWT sessions | 5 | 12 | 12 |
| Interval between ESWT sessions [days] | 7 | 3.5 | 3.5 |
| No. of shock waves per ESWT session | 3000 | 2000 | 1000 |
| Bar / EFD of shock waves | 3.5 / 0.12 mJ/mm<sup>2</sup> | 0.10 mJ/mm<sup>2</sup> | 0.01-0.16 mJ/mm<sup>2</sup> |
| Frequency of shock waves [Hz] | Not provided | 7 | 7 |
| Control treatment 1 |  |  |  |
| No. of subjects | 20 | 15 | 15 |
| Procedure | Single injection of 6 ml 1% trimecaine and 7 mg of betamethazone (C1) | Hyperthermia using hot packs, ultrasound and TENS (C) | Hyperthermia using hot packs (20 min), ultrasound (5 min) and TENS (15 min) (C) |
| Control treatment 2 |  |  |  |
| No. of subjects | 20 |  |  |
| Procedure | Radiofrequency Medial Branch Neurotomy (C2) |  |  |
| Follow-up interval | M2, M6 and M12 | Not provided | Not provided |
| Investigated variables | Pain (VAS score), Oswestry Disability Index (ODI) | Pain (VAS score), dynamic balance ability | Pain (VAS score), ODI, Beck depression index |
| Definition of treatment success | Not provided | Not provided | Not provided |
| VAS at baseline (ESWT) | Not provided | 7.2 ± 0.8 (SD or SEM) | 7.0 ± 0.76 (SD or SEM) |
| VAS at follow-up (ESWT) | Not provided | 4.5 ± 1.1 (SD or SEM) | 3.6 ± 1.1 (SD or SEM) |
| Outcome | C2 > rESWT > C1 | rESWT > C | fESWT > C |

| **Study** | **Moon et al. [35]** | **Notarnicola et al. [36]** | **Tomska et al. [37]** |
| --- | --- | --- | --- |
| Diagnosis | Sacroiliac joint pain | LBP | Back pain |
| Type of study | RCT | RCT | RCT |
| Power analysis | Not reported | Not reported | Not reported |
| No. of subjects | 15 | ? (total 30) | 36 |
| Device | Aries (Dornier MedTech, Wessling, Germany) | Minilith SL1 (Storz Medical) | Not reported |
| Type of ESWT | fESWT | fESWT | rESWT |
| Applicator | --- | --- | Not reported |
| No. of ESWT sessions | 1 | 3 | 6 |
| Interval between ESWT sessions [days] | N/a | 7 | 7 |
| No. of shock waves per ESWT session | 2000 | 2000 | 2000 |
| Bar / EFD of shock waves | 0.09-0.25 mJ/mm<sup>2</sup> (maximum tolerated by the patient) | 0.03 mJ/mm<sup>2</sup> | 60-100 mJ |
| Frequency of shock waves [Hz] | 3 | Not reported | 5-10 |
| Control treatment (C) |  |  |  |
| No. of subjects | 15 | ? (total 30) | 37 |
| Procedure | Sham ESWT | Individual exercise treatment | Deep electromagnetic stimulation |
| Follow-up interval | W1 and W4 | M1 and M3 | W0, W2 and W4 after treatment |
| Investigated variables | Pain (VAS score), ODI | Pain (VAS score), Roland Morris Disability Questionnaire, ODI | Pain (VAS score) |
| Definition of treatment success | Not provided | Not provided | Not provided |
| VAS at baseline (ESWT) | 6.42 ± ? | Not provided | 6.8 ± 2.3 (Mean ± SD) |
| VAS at follow-up (ESWT) | 3.64 ± ? | Not provided | 2.6 ± 2.0 (Mean ± SD) |
| Outcome | fESWT > C | ESWT > C | rESWT ≈ C |

| Study | Walewicz et al. [38] | Walewicz et al. [39] | Eftekharsadat et al. [40] |
| --- | --- | --- | --- |
| Diagnosis | Chronic LBP | Chronic LBP<br>(discopathy of the L5-S1 spine segment) | Inferior trigger points in the quadratus lumborum muscle |
| Type of study | RCT | RCT | RCT |
| Power analysis | Yes | Not reported | Yes |
| No. of subjects | 20 | 20 | 27 |
| Device | Pro-Shock Waves<br>(Cosmogamma, Indonesia) | Cellactor (Storz Medical) | enPulsPro (Zimmer MedizinSysteme, Neu-Ulm, Germany) |
| Type of ESWT | rESWT | rESWT | rESWT |
| Applicator | Not reported | Not reported | Not reported |
| No. of ESWT sessions | 10 | 10 | 5 |
| Interval between ESWT sessions [days] | 3.5 | 3.5 | 7 |
| No. of shock waves per ESWT session | 2000 | 2000 | 1500 |
| Bar / EFD of shock waves | 2.5 / 0.1 mJ/mm<sup>2</sup> | 2.5 / --- | 0.1 mJ/mm<sup>2</sup> |
| Frequency of shock waves [Hz] | 5 | 5 | 10-16 |
| Additional treatment | Functional training | Core stability treatment | Stretching exercises, education, acetaminophen |
| Control treatment (C) |  |  |  |
| No. of subjects | 20 | 20 | 27 |
| Procedure | Sham ESWT | Sham ESWT | Injection of corticosteroid |
| Follow-up interval | M1 and M3 | M1 and M3 | W2, W4 |
| Investigated variables | Pain (VAS score), Laitinen Pain Scale, ODI | Pain (Laitinen Pain Scale), Roland–Morris Questionnaire, original Schober Test, assessment of postural sway | Pain (VAS score), Pressure-pain threshold, ODI, Quality of Life |
| Definition of treatment success | Not provided | Not provided | Not provided |
| VAS at baseline (ESWT) | 4.7 ± 1.9 (Mean ± SD) | Not provided | 7.63 ± 0.27 (Mean ± SD) |
| VAS at follow-up (ESWT) | 2.0 ± 2.0 (Mean ± SD) | Not provided | 5.11 ± 0.36 (Mean ± SD) |
| Outcome | rESWT > C | rESWT > C | rESWT ≈ C |

| **Study** | **Kong et al. [41]** | **Celik et al. [42]** |
| --- | --- | --- |
| Diagnosis | Chronic LBP | Chronic LBP |
| Type of study | RCT | RCT |
| Power analysis | No | No |
| No. of subjects | 100 | 25 |
| Device | HK. ESWO-AJ<br>(Shenzhen Huikang<br>Medical Apparatus<br>Co., Ltd., Shenzhen,<br>China) | E1000 (EMD Medical<br>Technologies, Ankara,<br>Turkey) |
| Type of ESWT | fESWT | fESWT |
| Applicator | --- | --- |
| No. of ESWT sessions | 5-10 | 12 |
| Interval between<br>ESWT sessions [days] | Not provided | 3.5 |
| No. of shock waves<br>per ESWT session | 1200 | 1500 |
| Bar / EFD of shock<br>waves | 0.1-0.2 mJ/mm<sup>2</sup> | 0.12 mJ/mm<sup>2</sup> |
| Frequency of shock<br>waves [Hz] | Not provided | 2.5 |
| Control treatment (C) |  |  |
| No. of subjects | 100 | 25 |
| Procedure | Laser therapy | Sham ESWT |
| Follow-up interval | Not provided | W6, W12 |
| Investigated variables | Pain (VAS score),<br>Quality of life | Pain (VAS score),<br>ODI, Hospital Anxiety<br>and Depression Scale,<br>Short-form 36 |
| Definition of<br>treatment success | Not provided | Not provided |
| VAS at baseline<br>(ESWT) | 6.3 ± 1.1 (SD or SEM) | 5 (2-8) (median<br>(range)) |
| VAS at follow-up<br>(ESWT) | Not provided | 0 (0-7) (median<br>(range)) |
| Outcome | fESWT > C | ESWT > C |

### Appendix B

Studies listed in the PEDro database [55] in which the efficacy of non-pharmacological, conservative treatments in the management of low back pain (LBP) was compared with the efficacy of pharmacological treatments and the combination thereof.

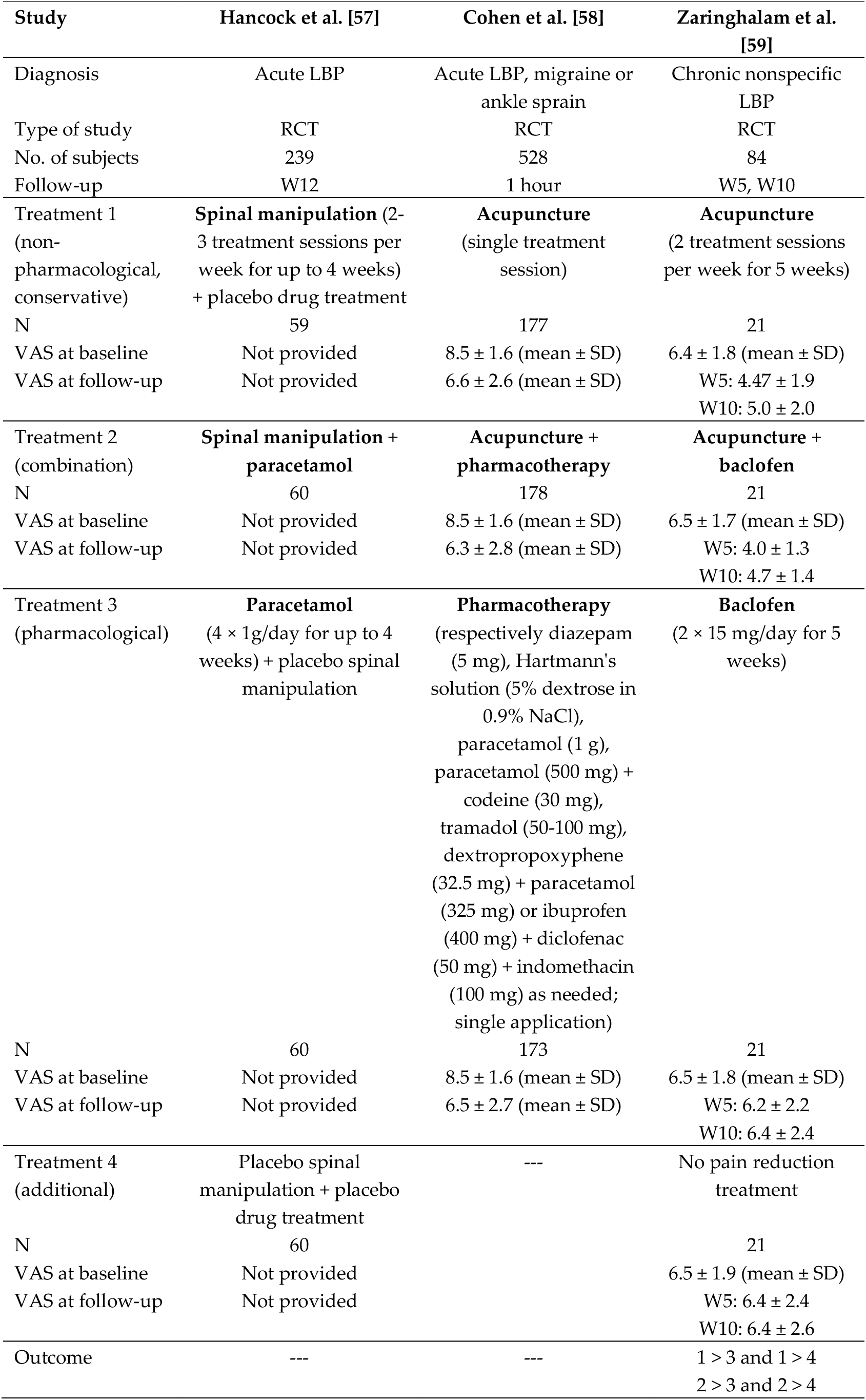

| Study | Hancock et al. [57] | Cohen et al. [58] | Zaringhalam et al. [59] |
| --- | --- | --- | --- |
| Diagnosis | Acute LBP | Acute LBP, migraine or ankle sprain | Chronic nonspecific LBP |
| Type of study | RCT | RCT | RCT |
| No. of subjects | 239 | 528 | 84 |
| Follow-up | W12 | 1 hour | W5, W10 |
| Treatment 1 (non-pharmacological, conservative) | **Spinal manipulation** (2-3 treatment sessions per week for up to 4 weeks) + placebo drug treatment | **Acupuncture** (single treatment session) | **Acupuncture** (2 treatment sessions per week for 5 weeks) |
| N | 59 | 177 | 21 |
| VAS at baseline | Not provided | 8.5 ± 1.6 (mean ± SD) | 6.4 ± 1.8 (mean ± SD) |
| VAS at follow-up | Not provided | 6.6 ± 2.6 (mean ± SD) | W5: 4.47 ± 1.9<br>W10: 5.0 ± 2.0 |
| Treatment 2 (combination) | **Spinal manipulation + paracetamol** | **Acupuncture + pharmacotherapy** | **Acupuncture + baclofen** |
| N | 60 | 178 | 21 |
| VAS at baseline | Not provided | 8.5 ± 1.6 (mean ± SD) | 6.5 ± 1.7 (mean ± SD) |
| VAS at follow-up | Not provided | 6.3 ± 2.8 (mean ± SD) | W5: 4.0 ± 1.3<br>W10: 4.7 ± 1.4 |
| Treatment 3 (pharmacological) | **Paracetamol** (4 × 1g/day for up to 4 weeks) + placebo spinal manipulation | **Pharmacotherapy** (respectively diazepam (5 mg), Hartmann's solution (5% dextrose in 0.9% NaCl), paracetamol (1 g), paracetamol (500 mg) + codeine (30 mg), tramadol (50-100 mg), dextropropoxyphene (32.5 mg) + paracetamol (325 mg) or ibuprofen (400 mg) + diclofenac (50 mg) + indomethacin (100 mg) as needed; single application) | **Baclofen** (2 × 15 mg/day for 5 weeks) |
| N | 60 | 173 | 21 |
| VAS at baseline | Not provided | 8.5 ± 1.6 (mean ± SD) | 6.5 ± 1.8 (mean ± SD) |
| VAS at follow-up | Not provided | 6.5 ± 2.7 (mean ± SD) | W5: 6.2 ± 2.2<br>W10: 6.4 ± 2.4 |
| Treatment 4 (additional) | Placebo spinal manipulation + placebo drug treatment | --- | No pain reduction treatment |
| N | 60 |  | 21 |
| VAS at baseline | Not provided |  | 6.5 ± 1.9 (mean ± SD) |
| VAS at follow-up | Not provided |  | W5: 6.4 ± 2.4<br>W10: 6.4 ± 2.6 |
| Outcome | --- | --- | 1 > 3 and 1 > 4<br>2 > 3 and 2 > 4 |

### Appendix C

Outcome of the present study of the subgroup of subjects with PSEQ score < 50 at baseline (*rESWT* group: n=16; *rESWT+C+E* group: n=21; *C+E* group: n=20) (these subjects could have reached the MCID of the PSEQ score as defined in the literature [44] when treating cnsLBP). The figure shows Tukey boxplots of (A) PSEQ score, (B) NRS score, (C) OLBPDQ score and (D) PHQ-9 score of subjects suffering from cnsLBP who were treated with respectively rESWT (dark gray bars), rESWT+C+E (light gray bars) or C+E alone (open bars) at baseline (X=0) and different follow-up times. The table summarizes key results of the statistical analysis of these data using two-way repeated measures ANOVA (P values < 0.05 are given boldface). Bonferroni’s multiple comparison test showed no statistically significant differences between groups.

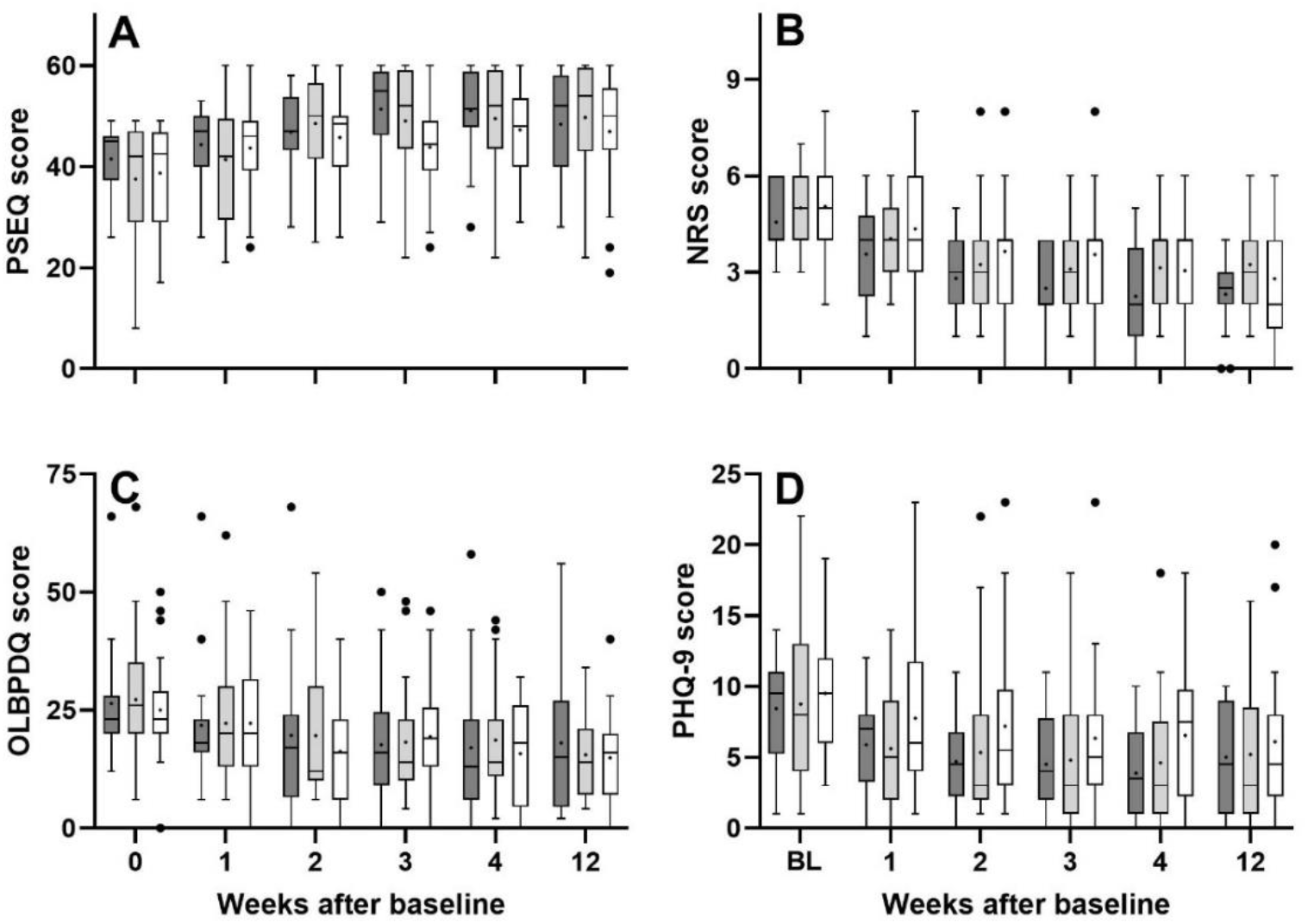

| ANOVA: source of variation | PSEQ score | NRS score | OLBDPQ score | PHQ-9 score |
| --- | --- | --- | --- | --- |
| Time | **<0.001** | **<0.001** | **<0.001** | **<0.001** |
| Treatment | 0.550 | 0.200 | 0.926 | 0.339 |
| Time × Treatment | 0.195 | 0.666 | 0.677 | 0.933 |
| Subject | **<0.001** | **<0.001** | **<0.001** | **<0.001** |

